# EpiLink: a simulation-based compatibility model for genomic transmission clustering in infectious disease surveillance

**DOI:** 10.64898/2026.06.16.26355814

**Authors:** Dominic Arthur, Christopher J. Banks, Rowland R. Kao

## Abstract

Identifying recently linked infections from pathogen genome sequences is central to infectious disease surveillance, yet many clustering approaches rely on fixed genetic distance thresholds whose relationship to transmission is often unclear. This limitation is especially important in rapidly growing outbreaks and superspreading events, where many cases may be sampled close together in time and share little genetic variation, making true transmission links difficult to distinguish from other closely related infections. Supervised models can improve discrimination, but they require labelled transmission data that are rarely available during outbreak response.

We developed EpiLink, a threshold-free method that estimates whether two cases are compatible with recent transmission. Here, compatibility means how well the observed genetic distance and sampling-time difference between two cases fit what would be expected if they were linked by defined recent transmission scenarios. EpiLink simulates plausible recent transmission histories while accounting for uncertainty in infection timing, testing delay, and mutation accumulation, then assigns higher scores to pairs whose observed differences are typical of those simulations.

EpiLink was evaluated using both synthetic and empirical SARS-CoV-2 outbreak data from the 2020 Boston epidemic. Two EpiLink variants were compared to a logistic regression model trained on labelled transmission data. One EpiLink variant assumed deterministic mutation accumulation, with genetic differences proportional to elapsed evolutionary time; the other accounted for stochasticity by sampling mutation counts from a Poisson distribution. The logistic regression model performed better at distinguishing linked from unlinked pairs, but EpiLink achieved comparable clustering accuracy. In the Boston data, EpiLink recovered clusters enriched for documented conference and skilled nursing facility outbreaks. EpiLink thus provides an interpretable, simulation-based approach for identifying recent transmission clusters when fixed thresholds are difficult to justify and labelled transmission data are unavailable.

**Author summary:** Grouping infectious disease cases into transmission clusters is a routine part of outbreak surveillance, but many methods rely on fixed genetic distance cut-offs that can be hard to interpret, especially when transmission is rapid and pathogen diversity is low. We developed EpiLink, which instead asks how consistent the observed genetic and sampling-time differences between two cases are with recent transmission. EpiLink simulates plausible transmission histories and scores each pair according to how typical its observed differences are within the simulated distributions. In simulated SARS-CoV-2 outbreaks, EpiLink nearly matched the clustering accuracy of a supervised model trained on labelled transmission pairs, without requiring labelled data. We found a practical trade-off: deterministic configurations performed best when model assumptions were well met, while configurations incorporating uncertainty were more robust when assumptions were misspecified. Applied to SARS-CoV-2 data from the 2020 Boston epidemic, EpiLink recovered clusters enriched for known outbreaks at a conference and skilled nursing facility. EpiLink offers a practical and interpretable approach for transmission clustering when labelled data are unavailable.

## Introduction

Identifying clusters of epidemiologically linked infections is a core task in infectious disease surveillance, underpinning outbreak investigation, situational awareness, and targeted public health intervention. Increasingly, this task is supported by pathogen genomic data, which can provide high-resolution information on relatedness between infections and has become an important component of outbreak detection and transmission analysis in public health surveillance [1, 2]. This task is particularly challenging in outbreaks shaped by superspreading events, in which a few infections give rise to a disproportionately large number of secondary cases [3]. In such settings, rapid bursts of transmission, incomplete sampling, and limited pathogen genetic diversity can make it difficult to distinguish true transmission links from coincidental similarity, especially when relying on sequence data alone [4, 5]. Superspreading has been documented across a range of pathogens, including SARS-CoV-1, MERS, Ebola, and SARS-CoV-2 [6–10], and is now recognised as a fundamental driver of transmission heterogeneity. Despite its epidemiological importance, the timely identification and characterisation of superspreading-driven molecular transmission clusters remain methodologically unresolved, particularly in the context of large-scale genomic surveillance.

Current analytical approaches often force a trade-off between detailed transmission reconstruction and simple threshold-based clustering. In many surveillance settings, however, the relevant question is narrower than full transmission-tree inference: given the observed genomes and sampling times for two cases, are they plausibly linked by recent transmission? This question is particularly salient in superspreading-driven outbreaks, where many cases may accumulate within a short time window and share minimal genetic divergence. Methods such as outbreaker2 [11, 12], TransPhylo [13], and SCOTTI [14] can explicitly model unsampled cases and uncertainty, yet they are generally best suited to detailed outbreak reconstruction and require assumptions and computational investment that may be difficult to support in real-time public health settings. By contrast, fixed-threshold and distance-based clustering approaches remain attractive because they are fast, scalable, and easy to implement [15–20].

Their practical appeal, however, comes at the cost of interpretability. Small genetic distances do not map cleanly onto recent transmission, particularly when pathogen diversity is low, mutation processes are stochastic, sampling is incomplete, or sampling times carry information that static genetic cut-offs ignore [4, 5, 16, 17, 20]. As a result, the epidemiological meaning of a pairwise link, or of a cluster derived from many such links, is often unclear. This limitation also applies to more flexible network-based strategies. Although hierarchical thresholding and community detection can reduce some of the instability associated with single cut-offs [21, 22], they still depend on underlying pairwise similarities whose relationship to recent transmission may be weak or ambiguous. The central challenge, therefore, is not only to detect structure in genomic data, but to do so in a way that yields clusters with a clear and defensible epidemiological interpretation.

A key part of this challenge is how pairwise genetic distances should be interpreted in biological and epidemiological terms when transmission is recent and highly clustered. Two sampled genomes separated by only a few single-nucleotide polymorphisms may suggest close relatedness, but they do not by themselves identify the exact relationship. Instead, that similarity may reflect direct infection, transmission through one or more unsampled intermediates, or infection from a common source followed by limited divergence across separate chains [23]. This ambiguity is especially pronounced for SARS-CoV-2 and other pathogens characterised by short generation intervals, rapid epidemic expansion, and relatively slow short-term sequence evolution [5, 17, 23–25]. In such settings, consensus genomes often remain identical or nearly identical across many epidemiologically distinct cases, so that genetic similarity alone provides only weak evidence about recent transmission at the scale relevant to superspreading events.

While the incorporation of temporal data can improve resolution, current methods often rely on fixed cut-offs [26] or supervised models trained on labelled transmission pairs [19], and therefore lack a principled way to carry uncertainty in timing, infectiousness, and mutation processes through to cluster inference.

To address this interpretability gap, we developed EpiLink, a threshold-free, simulation-based method for evaluating recent-transmission compatibility between sampled cases. EpiLink does not seek to infer a fully resolved transmission tree. Instead, it assesses whether the observed temporal separation and genetic distance between two cases are consistent with Monte Carlo draws generated under a user-defined set of recent-transmission scenarios, such as direct transmission between two sampled cases or infection through a shared unsampled ancestor. This formulation is designed for the operational scale at which surveillance often functions, where the aim is not necessarily to identify an exact infector, but to determine whether two cases plausibly belong to the same recent transmission neighbourhood. By explicitly modelling uncertainty in infection timing, infectiousness, unsampled transmission, and mutation accumulation, EpiLink produces pairwise compatibility scores with a clear mechanistic interpretation, while avoiding reliance on arbitrary distance thresholds or labelled training data.

In this study, we evaluate EpiLink using both synthetic outbreak data and a published SARS-CoV-2 dataset from Boston [27]. We first examine the recent-transmission compatibility surface induced by the model across combinations of temporal and genetic distance, and assess the extent to which graph sparsification can reduce network density while preserving informative pairwise structure. We then evaluate pairwise discrimination and graph-based cluster recovery under baseline conditions and systematic perturbations of key simulation parameters, and assess the stability of inferred clusters as surveillance data accumulate. Finally, we test whether the method recovers epidemiologically documented superspreading structure in the Boston dataset. Throughout, we compare EpiLink with logistic regression trained on labelled pairs as a supervised reference, and include TreeCluster as a supplementary phylogenetic hard-clustering comparator.

## Materials and methods

### EpiLink compatibility model

EpiLink is a pairwise compatibility model that assesses whether the observed consensus genetic distance *g*_ij_ and sampling-time difference *t*_ij_ between two sampled cases (*i, j*) are consistent with one or more candidate latent transmission histories (Fig 1). Rather than inferring a unique transmission tree, EpiLink evaluates compatibility with hypothesised epidemiological relationships while accounting for uncertainty in infection timing, testing delay, and mutation accumulation.

**Fig 1.**
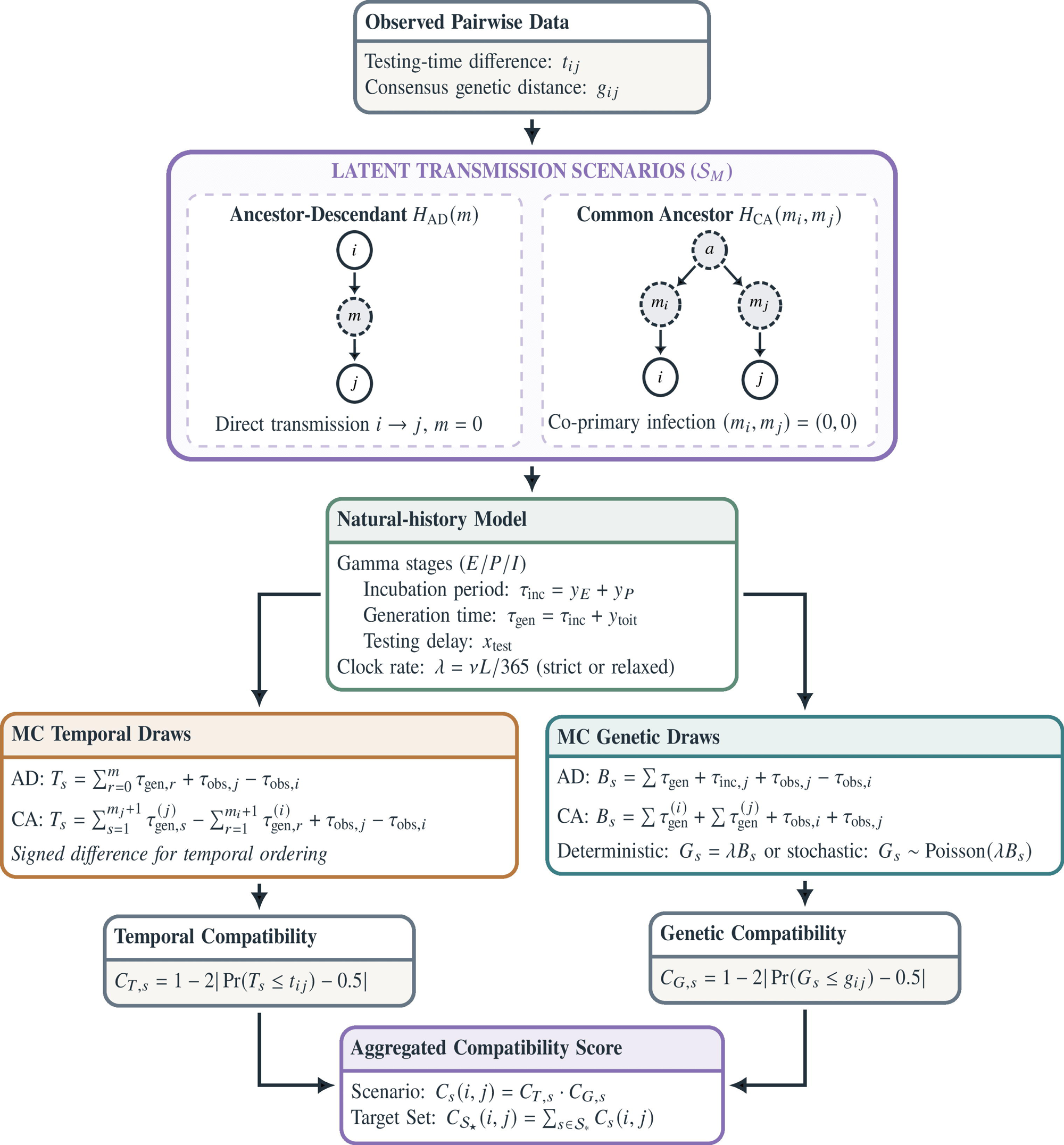
Overview of the EpiLink compatibility model. For a sampled pair (*i, j*), observed testing-time difference *t*_ij_ and consensus genetic distance *g*_ij_ are evaluated against Monte Carlo distributions generated under latent transmission scenarios S_M_ (ancestor-descendant *H*_AD_ and common-ancestor *H*_CA_ histories). A shared natural-history model drives parallel temporal and genetic compatibility tracks; scenario-level scores *C*_s_(*i, j*) = *C*_T,s_(*i, j*) *C*_G,s_(*i, j*) are summed over the target subset S_*_ to yield the final pairwise score *C*_S*_ (*i, j*). Full notation is defined in the main text.

We considered two classes of latent transmission scenarios up to a maximum hidden depth *M* :

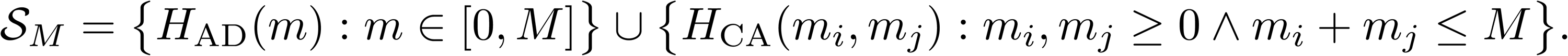

Here, *H*_AD_(*m*) denotes an ancestor-descendant scenario in which case *j* descends from case *i* through *m* unsampled intermediates, and *H*_CA_(*m*_i_*, m*_j_) denotes a common-ancestor scenario in which both cases descend from a shared unsampled source independently. The cases *H*_AD_(0) and *H*_CA_(0, 0) correspond to direct transmission and co-primary infection, respectively. In the default configuration used throughout this study, the target subset is S_*_ = {*H*_AD_(0)*, H*_CA_(0, 0)}, implying a maximum depth of *M* = 0.

For each scenario *s* ∈ S_M_, EpiLink generates Monte Carlo draws from a shared natural-history model with Gamma-distributed latent (*E*), presymptomatic infectious (*P*), and symptomatic infectious (*I*) stage durations (*y*_E_, *y*_P_, and *y*_I_ denote random variables), together with a Gamma-distributed delay from symptom onset to testing or sampling (*x*_test_) [28–30]. Writing the incubation period as *τ*_inc_ = *y*_E_ + *y*_P_ and the generation time as *τ*_gen_ = *τ*_inc_ + *y*_toit_ (where *y*_toit_ is the time from onset of infectiousness to transmission), the total observation interval from exposure to testing is *τ*_obs_ = *τ*_inc_ + *x*_test_; these draws induce a scenario-specific distribution for the expected testing-time difference *T*_s_. The same latent history defines an effective transmission-related branch length *B*_s_: under ancestor-descendant scenarios, this is the sum of generation intervals along the chain; under common-ancestor scenarios, it is the sum of lineage lengths from the shared source to both sampled descendants. The full derivation of *T*_s_ and *B*_s_ is given in S1 Text.

Branch-length draws were mapped to expected genetic distances using a per-genome daily substitution rate *λ* = *νL/*365, where *ν* is the per-site per-year substitution rate and *L* is the genome length. Under a deterministic clock, *G*_s_ = *λB*_s_; under a stochastic mutation process, *G*_s_ | *B*_s_ ∼ Poisson(*λB*_s_) [31]. A relaxed uncorrelated log-normal clock draws *ν* from a log-normal distribution [32].

Compatibility was quantified by locating each observed value within the corresponding scenario-specific empirical distribution. For observed value *x*_obs_ and *n*_s_ Monte Carlo draws from scenario *s*:

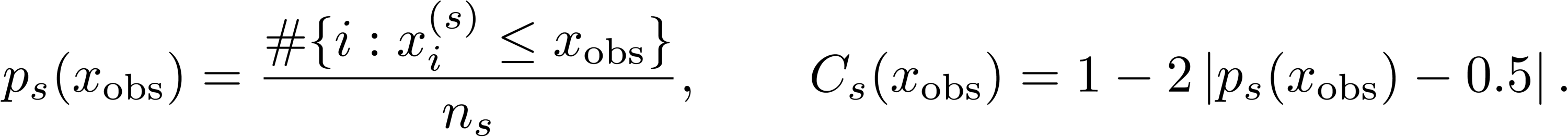

Temporal and genetic compatibilities *C*_T,s_(*i, j*) and *C*_G,s_(*i, j*) are combined as their product to obtain the scenario-level score *C*_s_(*i, j*) = *C*_T,s_(*i, j*) *C*_G,s_(*i, j*), and then summed across the target subset:

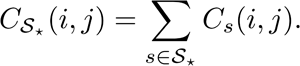

This is a compatibility model rather than a calibrated probabilistic transmission model: scores reflect how centrally the observed pair sits within the simulated distributions induced by S_*_, not posterior transmission probabilities.

### Synthetic benchmark dataset

We evaluated EpiLink using synthetic outbreak datasets derived from SCoVMod—a spatially explicit agent-based model that simulates transmission across Scotland using demographic structure, mobility patterns, and time-varying contact behaviour [33].

Starting from SCoVMod infection-history and transmission-event outputs, we constructed a directed transmission network, resolved multiply infected nodes by retaining the earliest recorded parent, and selected a weakly connected component containing approximately 5,000 nodes. A rooted arborescence was then obtained by computing the maximum spanning arborescence with edge weights defined as the negative simulation timestep, thereby favouring earlier parental assignments. The resulting transmission tree comprised 4,990 nodes and 4,989 directed edges.

Transmission heterogeneity in the underlying SCoVMod outbreak was summarised by fitting a negative-binomial offspring distribution to the full transmission network, yielding an estimated dispersion parameter of *k*^^^ = 0.233 (95%CI: 0.232–0.235) and mean reproductive number *R*^̄^_t_ ≈ 1.00 (95% CI: 0.992–1.006). Approximately 15.6% of individuals accounted for 80% of onward transmission, consistent with the superspreading dynamics motivating this work. Synthetic epidemic dates and pairwise genetic distances were generated by overlaying the EpiLink natural-history model onto this fixed transmission tree topology.

### Baseline epidemiological and evolutionary parameters

Key baseline evaluation parameters (Table 1) were drawn from published SARS-CoV-2 estimates [28, 34, 35]. Other *E/P/I* infectiousness profile parameters were kept fixed at their fitted values (see S1 Text). To aid interpretation, the mean and coefficient of variation (CV) of the Gamma distributions are shown; in implementation, these are mapped to the corresponding shape *k* and scale *θ* parameters, such that increasing the CV decreases the shape parameter and increases the scale parameter. Multipliers of {0.75, 1.25} were applied to strictly positive parameters (incubation mean and CV, testing-delay mean and CV, and substitution rate), whereas multipliers of {0.0, 1.25} were applied to the clock-relaxation parameter, allowing inclusion of a strict-clock case. In total, this produced 12 perturbation scenarios.

**Table 1.** Baseline evaluation parameters and perturbation design.

| Parameter | Baseline value | Lower mult. | Upper mult. |
| --- | --- | --- | --- |
| Incubation mean | 5.505 days | 0.75 | 1.25 |
| Incubation CV | 0.415 | 0.75 | 1.25 |
| Testing-delay mean | 1.0 day | 0.75 | 1.25 |
| Testing-delay CV | 1.0 | 0.75 | 1.25 |
| Substitution rate | $10^{-3}$ sub/site/year | 0.75 | 1.25 |
| Clock relaxation | 0.33 | 0.0 | 1.25 |
| *Fixed simulation parameters:* |  |  |  |
| Synthetic genome length | 5,000 bp | — | — |
| Monte Carlo draws | 10,000 | — | — |
| Target subset | $\{H_{AD}(0), H_{CA}(0, 0)\}$ | — | — |
| Sparsification threshold | $10^{-4}$ | — | — |

### Model variants

Synthetic pairwise temporal and genetic data—either deterministic (D) or stochastic (S)—were supplied as input to EpiLink, which applies either a deterministic or stochastic formulation for compatibility score inference. This yields four EpiLink variants: EDD (deterministic inference, deterministic input), EDS (deterministic inference, stochastic input), ESD (stochastic inference, deterministic input), and ESS (stochastic inference, stochastic input). Two logistic regression models (LD and LS) were fitted as supervised reference comparators using the same pairwise features supplied to EpiLink. Both were trained on a random 10% subsample of all case pairs, ensuring they had access to the same observables as EpiLink but with the addition of transmission labels.

### Simulation design and sensitivity analysis

We conducted a simulation study to evaluate inference performance and robustness under a range of epidemiological and evolutionary conditions. Synthetic outbreak datasets were generated on the fixed 4,990-case transmission tree under the baseline parameterisation described above. Sensitivity analyses were designed to assess the effect of perturbing the two core mechanisms governing pairwise compatibility: (1) the infection-to-sampling interval, varied through the mean and CV of the incubation-period and testing-delay distributions; and (2) the molecular clock rate, varied through the substitution rate and degree of relaxation. Perturbations were applied one at a time.

Each scenario was evaluated under two inference conditions. In the **matched** condition, inference parameters were updated to match the perturbed data-generating values, representing best-case performance under each altered regime. In the **mismatched** condition, data-generation parameters were perturbed, but inference parameters were held fixed at their baseline values, quantifying robustness to parameter misspecification. The logistic regression comparators were treated analogously: retrained for each perturbation scenario under the matched condition, and trained once on baseline data only under the mismatched condition. Performance losses were calculated as relative fractional changes from the unperturbed matched baseline.

### Sparsification analysis

We identified minimum score thresholds at which nearly all total edge weight was retained while graph density was substantially reduced. For each model, the optimal sparsification threshold was defined as the smallest value in {10^−4^, 10^−3^, 10^−2^, 10^−1^} that retained at least 99.5% of total edge weight.

### Graph construction and community detection

For all synthetic experiments, weighted graphs were constructed from pairwise compatibility scores (or logistic probabilities) after excluding edges with weights below the sparsification threshold (10^−4^). Community detection was performed using the Leiden algorithm [36] with a modularity-based objective. The resolution parameter was swept over *γ* ∈ {0.1, 0.2*, . . .,* 1.0} with 10 random restarts at each value, and the resolution yielding the highest BCubed F1 score was selected for each model.

### Performance metrics

Clustering performance was assessed using BCubed precision and recall [37] against overlapping reference neighbourhoods derived from the transmission tree. For each node *u*, the reference cluster consisted of *u* together with all of its direct infectees. BCubed precision measures, for each case, the proportion of inferred co-cluster members that truly share its reference neighbourhood; BCubed recall measures the proportion of the reference neighbourhood recovered in the inferred cluster; BCubed F1 is their harmonic mean, averaged across all cases.

Pairwise discrimination was quantified using average precision (AP). Positive pairs were rare, comprising approximately 0.14% of all sampled pairs, making precision-recall summaries more informative than ROC-based measures in this highly imbalanced setting [38]. A pair was labelled positive if the two cases were connected by direct transmission or were sibling infectees sharing the same parent in the transmission tree.

### Temporal stability analysis

Temporal stability was evaluated by grouping sampled cases into weekly availability bins and recomputing clustering after each cumulative weekly update using only cases available up to that time point. Consecutive weekly partitions were compared using three measures: forward overlap (the fraction of a later cluster recovered from its best-matched earlier cluster), backward overlap (the fraction of an earlier cluster retained in its best-matched later cluster), and the Jaccard index, which provides a symmetric stability measure that penalises both splitting and merging. For the logistic regression comparators, training labels were restricted to the first three cumulative weekly bins (86 initial cases), reflecting a limited early-supervision setting. The clustering resolution for all models was fixed using the early window and held constant across subsequent cumulative analyses.

### Boston empirical dataset and clustering

The empirical evaluation used 772 high-quality SARS-CoV-2 whole-genome sequences (*>*98% coverage) collected by Massachusetts General Hospital (MGH) and the Massachusetts Department of Public Health (DPH) during the first wave of the COVID-19 pandemic in Boston, Massachusetts (March to May 2020) [27]. Sequences were aligned using Nextclade against the SARS-CoV-2 reference genome (GenBank accession no. MN908947.3) [39]. Pairwise consensus SNP distances were computed under the TN93 substitution model [40]. Sequence metadata included collection date and an exposure category label (Conference, SNF, BHCHP, City, Other) derived from epidemiological investigation. EpiLink scores were computed for all 297,606 unique pairs using the stochastic variant under baseline inference parameters. A weighted graph was constructed with minimum edge weight 10^−4^, and the Leiden algorithm was run at resolution *γ* = 0.3 with 10 restarts. Clusters of size ≥ 2 were retained. Focus clusters were those containing at least one Conference- or SNF-linked case; deviation from background exposure prevalence was tested using a *χ*^2^ goodness-of-fit test.

### TreeCluster comparison

As a supplementary tree-based hard-clustering comparator, we evaluated TreeCluster [41] on time-calibrated phylogenies. For the synthetic benchmark, distance-based trees were inferred separately from deterministic and stochastic pairwise genetic distance matrices and calibrated with TreeTime [42] using the simulated sampling dates. For the Boston dataset, a maximum-likelihood tree was inferred from the aligned genomes using IQ-TREE 3 with model selection and fast search options [43], then calibrated with TreeTime using collection dates. TreeCluster was run on dated Newick trees with thresholds expressed as calendar days divided by 365. In the synthetic benchmark, the max clade, avg clade, and single linkage modes were swept over thresholds from 7 to 70 days in 7-day increments and scored using BCubed F1 against the same overlapping SCoVMod reference memberships. For the Boston dataset, avg clade was swept over the same threshold grid and compared with EpiLink-Leiden partitions using adjusted Rand index (ARI) and adjusted mutual information (AMI); the exported Boston TreeCluster partition used avg clade at 28 days. TreeCluster returns hard partitions rather than pairwise linkage scores, so AP and related ranking metrics were not calculated.

### Software and data availability

The EpiLink software is available under an MIT licence at https://github.com/ydnkka/epilink and is archived on Zenodo (DOI: 10.5281/zenodo.20402076). All code, configuration files, and derived outputs required to reproduce the analyses presented in this manuscript are available in the evaluation branch at https://github.com/ydnkka/epilink/tree/evaluation, also archived on Zenodo (DOI: 10.5281/zenodo.20546213). The Boston empirical sequences and metadata were derived from the previously published dataset [27]. The synthetic benchmark was derived from SCoVMod outputs described in Banks et al. [33].

### Ethics

The SARS-CoV-2 genomic sequences and epidemiological metadata analysed here were collected and anonymised by Massachusetts General Hospital and the Massachusetts Department of Public Health as part of the study reported by Lemieux et al. [27], which received approval from the MGH Institutional Review Board. The present work constitutes a secondary analysis of that published dataset and did not involve the collection of new human data or biological specimens; no additional ethics approval was required.

## Results

### EpiLink produces interpretable compatibility surfaces

Compatibility scores were concentrated among short sampling gaps and low-to-moderate SNP distances, with scores falling for pairs far from the temporal-genetic ranges implied by the target scenarios (Fig 2). Two broad high-compatibility regimes emerged: one at very short temporal gaps and modest genetic distance, consistent with co-primary infection, and one at slightly longer gaps but minimal genetic distance, consistent with direct transmission. The stochastic formulation (Fig 2B) produced broader, less sharply bounded regions than the deterministic (Fig 2A). EpiLink thus assigns greatest support to pairs whose observed distances are plausibly explained by either scenario, while strongly down-weighting pairs outside these ranges.

**Fig 2.**
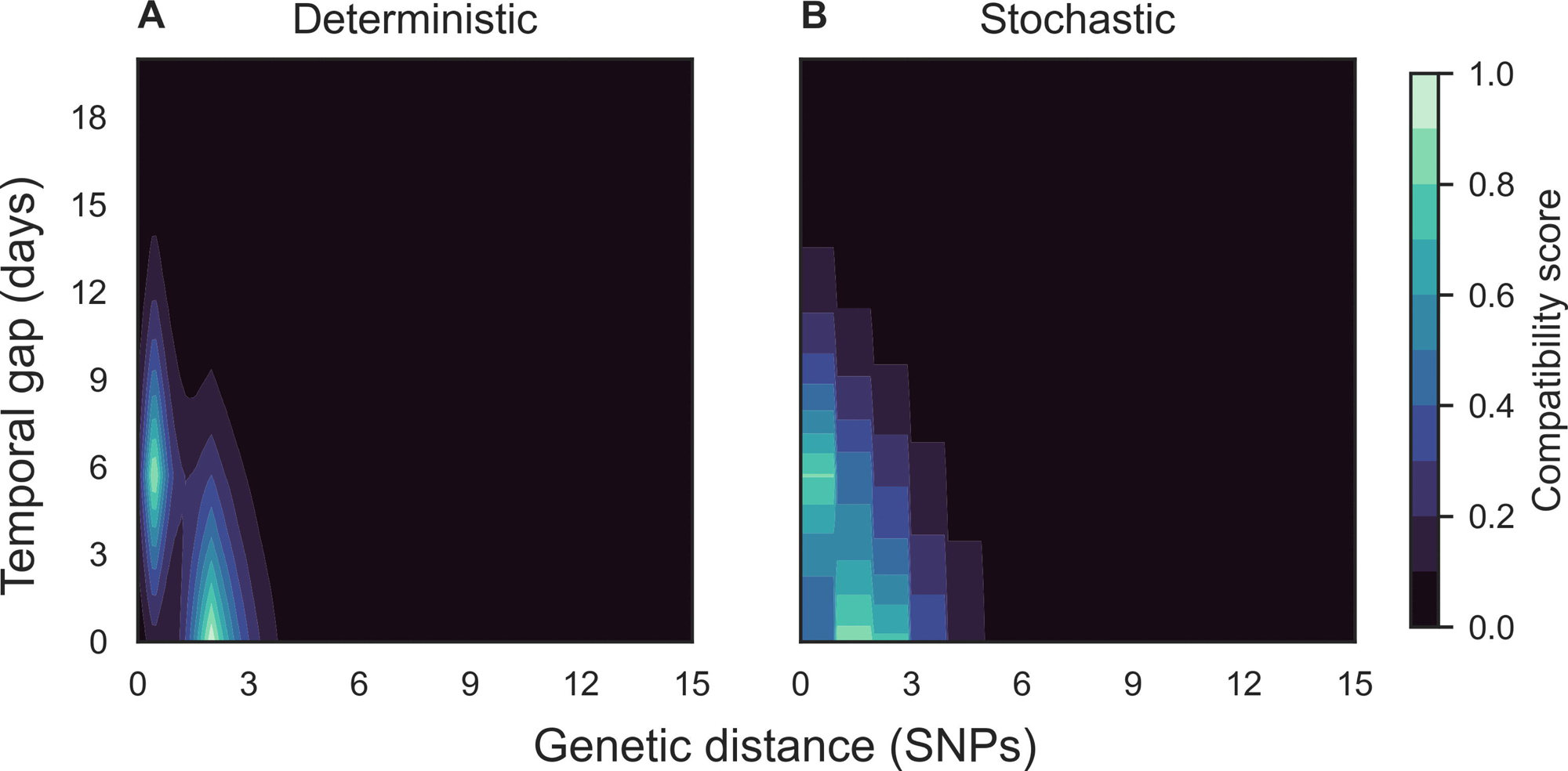
EpiLink compatibility surfaces. Contour plots of EpiLink compatibility as a function of genetic distance and temporal gap, using baseline inference parameters. (A) deterministic genetic component; (B) stochastic genetic component.

### ESD approaches supervised clustering accuracy, with partitions stabilising as surveillance accumulates

Our evaluation used a 4,990-node transmission tree yielding 12,447,555 scored pairs, of which approximately 0.145% were positive. We identified 10^−4^ as a reasonable sparsification threshold: at this cut-off, 87–97% of edges were removed yet more than 99.8% of total graph weight was preserved, reducing pipeline runtime from 18.2–26.8 seconds to 0.68–2.60 seconds per model (10–39× speed-up). We therefore applied this threshold in all subsequent clustering analyses.

In the baseline scenario, all six models substantially exceeded chance discrimination, with relative APs ranging from 63× to 393× (Table 2; Fig 3). LD achieved the highest pairwise discrimination and ESD the strongest among EpiLink variants, with ESD closely matching LD in clustering (F1 = 0.644 vs. 0.647), indicating that the AP advantage of the supervised model did not translate proportionally into cluster-level differences. Deterministic-input models (LD/ESD/EDD) consistently outperformed their stochastic-input counterparts (LS/ESS/EDS) at both discrimination and clustering. Yet these stochastic-input models still achieved moderately coherent partitions despite weak pairwise discrimination, reinforcing that aggregating scores can recover meaningful transmission structure even when individual scores are noisy.

**Fig 3.**
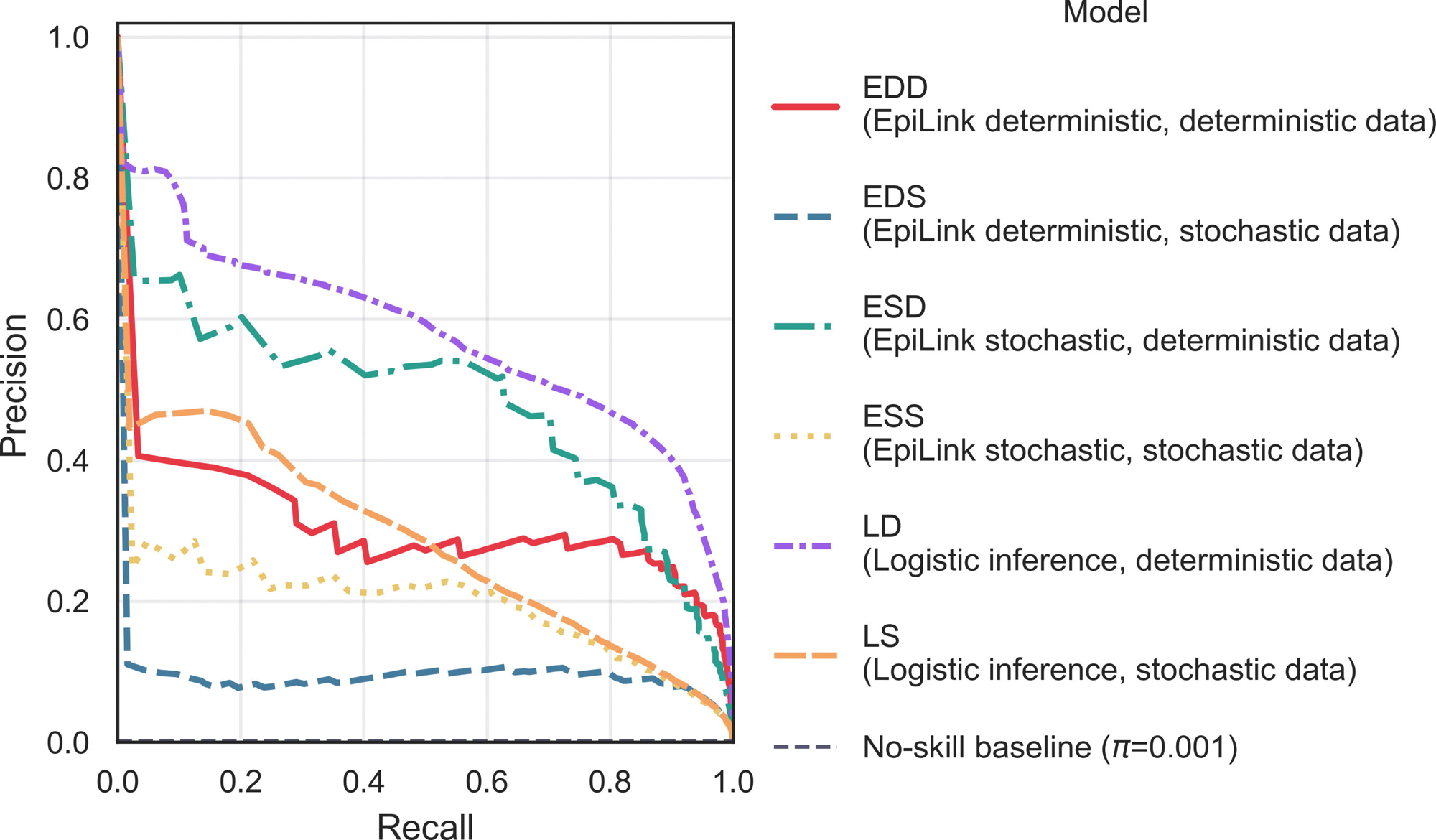
Baseline synthetic performance across all six models. Precision-recall analysis examining the informativeness of pairwise weights—EpiLink scores and logit probabilities. Performance is summarised with average precision (AP) with 95% confidence interval.

**Table 2.** Baseline performance metrics for all six models. AP is the average precision with 95% confidence interval; F1 is the BCubed F1 score; Relative AP is the AP relative to positive class prevalence (0.00145); Partition stability is the mean F1 between consecutive Leiden resolution steps (measures robustness to resolution choice); Temporal stability is the mean Jaccard index between consecutive weekly partitions.

| Model | AP (95% CI) | Relative AP | F1 | Partition stability | Temporal stability |
| --- | --- | --- | --- | --- | --- |
| LD | 0.568 (0.560–0.576) | 393 | 0.647 | 0.895 | 0.888 |
| ESD | 0.469 (0.462–0.477) | 324 | 0.644 | 0.851 | 0.893 |
| EDD | 0.300 (0.295–0.305) | 207 | 0.629 | 0.871 | 0.916 |
| LS | 0.275 (0.271–0.281) | 190 | 0.572 | 0.927 | 0.926 |
| ESS | 0.190 (0.187–0.194) | 132 | 0.559 | 0.830 | 0.825 |
| EDS | 0.090 (0.089–0.092) | 63 | 0.495 | 0.834 | 0.853 |

Cluster assignments became more stable after the first few weekly updates, although the level and timing of stabilisation varied by model (Fig. S1 Fig).

As a supplementary comparison with dated-tree hard clustering, TreeCluster achieved F1 scores of 0.590 on stochastic synthetic data (avg clade, 21 days) and 0.588 on deterministic synthetic data (max clade, 28 days) (Fig. S2 Fig). These values were below LD, ESD, and EDD, but above LS, ESS, and EDS, placing TreeCluster between the strongest deterministic-input models and the weaker stochastic-input comparators. Because TreeCluster returns hard partitions rather than pairwise scores, AP was not computed.

### Shorter incubation period and reduced clock produce the largest losses

The baseline performance ordering was largely preserved across all twelve perturbation scenarios (Fig. S3 Fig). Sensitivity analyses showed that AP (S4 Fig) was more responsive to perturbation than F1 (Fig 4), indicating that ranking performance was more vulnerable to misspecification than cluster-level accuracy. The most consistently degrading scenarios across both matched and mismatched conditions were reduced substitution rate and shortened incubation mean, with losses generally larger under mismatch. Increased incubation CV produced a markedly asymmetric pattern: under matched conditions most models improved, with EDS gaining the most in relative terms (F1 +25.2%; AP +253%, corresponding to absolute gains of +0.125 and +0.229); under mismatched conditions, deterministic-input models suffered substantial relative F1 losses (LD -25.4%, ESD -23.1%, EDD -21.6%) while stochastic-input models were largely unaffected (EDS +0.4%). Testing-delay perturbations produced negligible relative F1 changes throughout. Among models, EDD showed the largest losses under reduced substitution rate and shortened incubation mean; EDS was the most robust EpiLink variant across scenarios by relative F1 change. Several perturbations improved performance, particularly longer incubation mean, reduced incubation CV, higher substitution rate, and removal of clock relaxation (strict clock), with gains most pronounced for EDD in F1 and EDS in AP.

**Fig 4.**
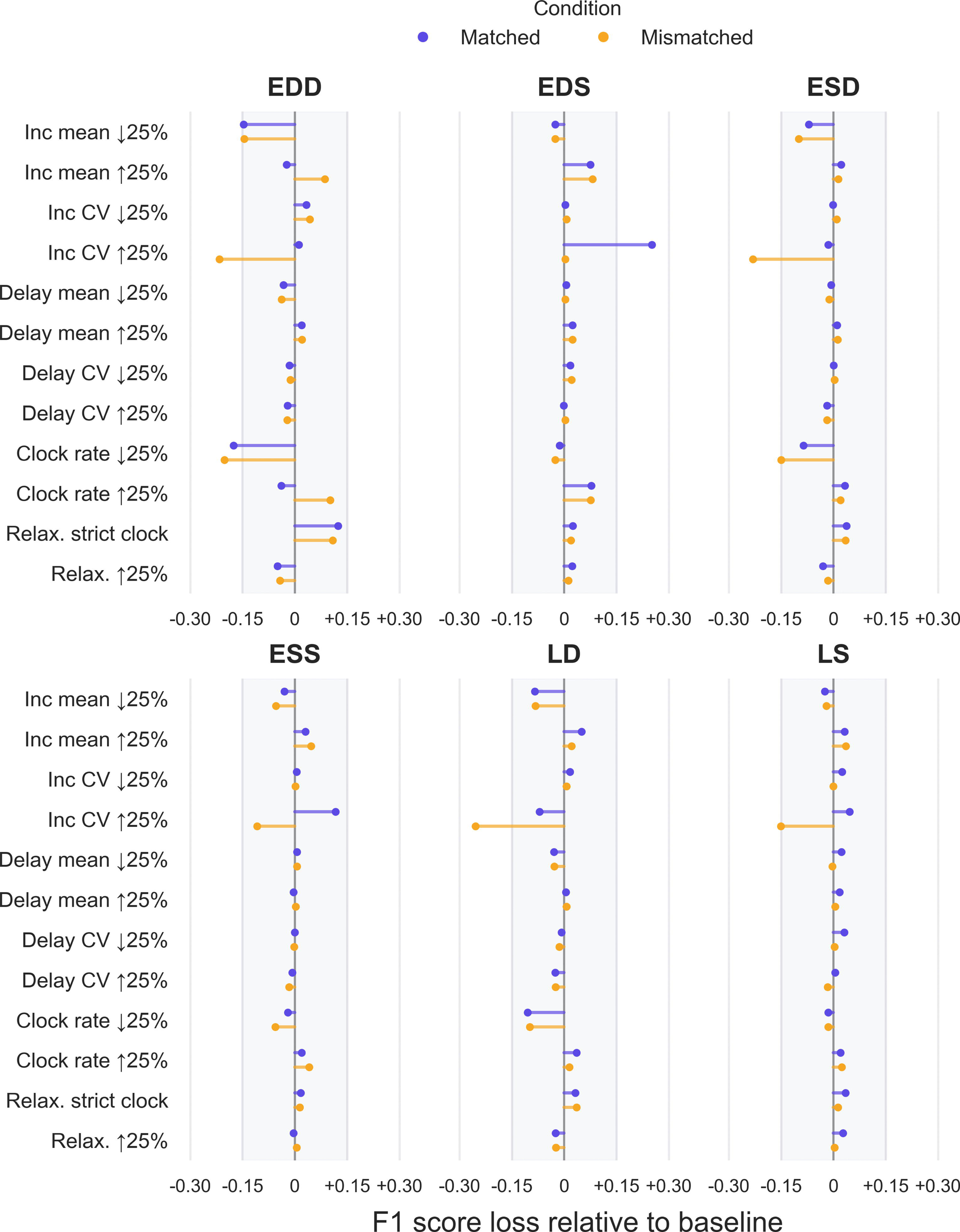
Sensitivity of F1 performance to perturbations of data-generating parameters. Lollipop plot showing relative best-F1 score loss compared with the unperturbed baseline for each model across twelve perturbation scenarios, under matched and mismatched inference conditions. Matched: inference parameters updated to reflect each perturbed scenario; mismatched: inference parameters held fixed at baseline. Each row corresponds to a perturbation scenario. Positive values indicate improved performance relative to baseline; negative values indicate degradation.

### Application to empirical data recovers exposure-enriched clusters

Applying EpiLink to 297,606 unique pairs from 772 Boston sequences yielded 77 clusters of size ≥ 2, containing 642 sequences in total, with a highly right-skewed size distribution (median = 3, mean = 8.3, max = 84) (Fig 5A). Five clusters containing at least one Conference- or SNF-linked case showed exposure compositions that differed significantly from background in all five (*χ*^2^, all *p <* 0.003) (Fig 5B).

**Fig 5.**
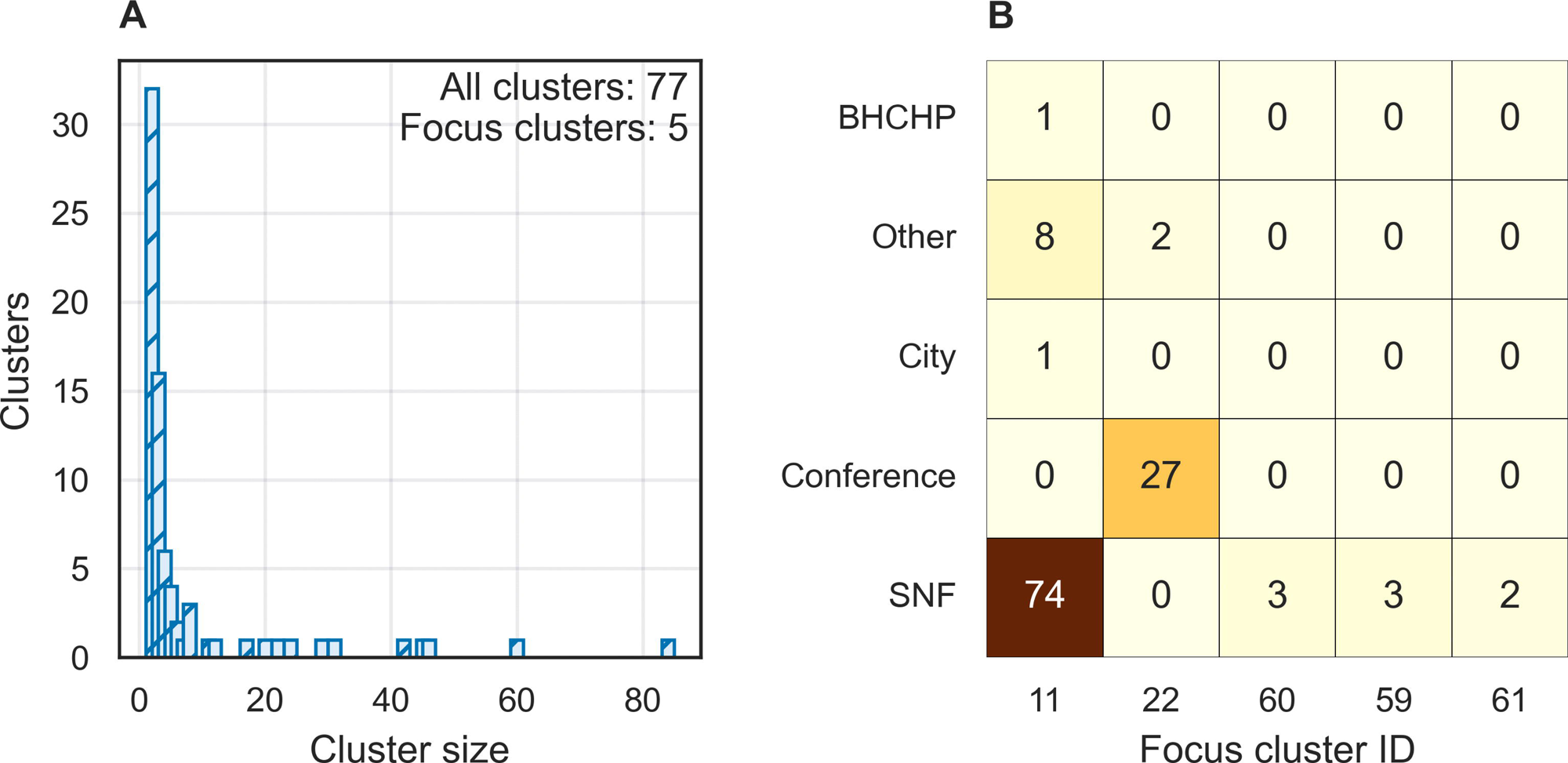
EpiLink clustering of the Boston MGH/DPH SARS-CoV-2 dataset. (A) Distribution of Leiden cluster sizes across all 772 sequences; 5 of 77 clusters contained at least one Conference- or SNF-linked case. (B) Heatmap of exposure category counts across these 5 clusters, annotated with raw case counts.

The two largest clusters containing known outbreak-associated cases illustrate the signal EpiLink recovers. Cluster 11 (*n* = 84) was dominated by SNF-linked cases (88.1%), with mean intra-cluster SNP distance of 0.69 (max 4) and mean sampling gap of 2.30 days (max 6) — consistent with a tight, contemporaneous outbreak. Cluster 22 (*n* = 29) was enriched for Conference-linked cases (93.1%), with similarly compact genetic and temporal structure (mean SNP distance 1.14, max 5; mean gap 1.43 days, max 7). In both cases, mean inter-cluster SNP distance was *>* 3, indicating higher average genetic separation from background while allowing occasional zero-distance matches outside the cluster. These results demonstrate that EpiLink recovers epidemiologically coherent clusters in real-world surveillance data, with cluster membership reflecting known exposure structure rather than sampling artefact.

The supplementary TreeCluster analysis of the Boston dataset showed a similar but more fragmented exposure-associated signal. TreeCluster with avg clade at 28 days yielded 111 clusters of size ≥ 2, containing 542 sequences, with 230 singletons. Fifteen TreeCluster clusters contained at least one Conference- or SNF-linked case. The largest SNF-associated cluster contained 71 sequences, including 69 SNF-linked cases, while the largest Conference-associated cluster contained 12 sequences, including 7

Conference-linked cases. Compared with EpiLink, TreeCluster assigned fewer samples to non-singleton clusters and split the exposure-associated signal across more clusters. Agreement between EpiLink and TreeCluster was moderate, with maximum ARI 0.553 at the selected 28-day TreeCluster threshold and Leiden resolution 0.5; at the same threshold, AMI reached 0.582 at Leiden resolution 0.4 (Fig. S5 Fig).

## Discussion

This study introduces EpiLink as an interpretable, label-free alternative to threshold-based genomic clustering, replacing hard cut-offs with a graded measure of recent-transmission compatibility that makes its biological assumptions explicit. The results clarify both where this approach adds value and where its limitations lie.

The motivation for EpiLink arose partly from the challenge of superspreading events, in which rapid, clustered transmission produces many cases with minimal genetic divergence within a short time window, making it difficult to distinguish true transmission links from coincidental similarity on sequence data alone. The Boston application illustrates that this is tractable: EpiLink recovered clusters strongly enriched for the skilled nursing facility and conference outbreaks without any supervised training, and cluster membership reflected documented exposure structure rather than sampling artefact [27]. That result is precisely the operational context for which the method was designed—consensus genomes and case times available, transmission labels absent, and the goal a recoverable recent-transmission neighbourhood rather than a direction-resolved transmission tree.

The synthetic benchmark places that result in context. Under matched baseline conditions, the supervised logistic comparator LD achieved the strongest pairwise discrimination. At the clustering level, however, ESD produced near-identical F1 performance to LD (0.644 vs. 0.647), indicating that the best EpiLink variant can approach supervised cluster-recovery accuracy when model assumptions are well-matched. This is consistent with what one expects: a model trained on labelled pairs has direct access to transmission information EpiLink does not, yet the clustering gap narrowed considerably. The more relevant comparison is between EpiLink and the threshold-based approaches it is intended to replace. The supplementary TreeCluster benchmark addresses this directly: TreeCluster achieved intermediate F1 performance, below the strongest EpiLink and logistic configurations but above the weaker stochastic-input comparators. In the Boston analysis, TreeCluster recovered exposure-associated structure but fragmented it into more, smaller clusters than EpiLink. This supports the central interpretation that EpiLink and dated-tree thresholding recover related signals, while EpiLink provides a graded pairwise compatibility score with an explicit recent-linkage interpretation before clustering. Within the EpiLink variants, the heterogeneity across EDD, EDS, ESD, and ESS is informative in its own right: each captures a distinct form of temporal-genetic model mismatch, and deterministic-input models consistently outperformed stochastic-input counterparts at both pairwise discrimination and cluster recovery. This indicates that the additional variance introduced by stochastic data generation degrades identifiability even when inference parameters are otherwise well-matched.

Temporal stability results support use in a rolling surveillance setting. Once cumulative case counts grew beyond the earliest weekly updates, cluster assignments were largely preserved across most consecutive updates, with EDD having the highest mean EpiLink Jaccard stability and ESD the highest minimum Jaccard stability. The early instability is an artefact of sparse case accumulation rather than intrinsic model fragility, and the asymmetry between forward and backward overlap reflects the expected structure of case accumulation: new cases enter existing clusters (high backward overlap) before those clusters are fully resolved.

The sensitivity analysis identifies two primary practical vulnerabilities: reduced substitution rate and shortened incubation period mean. These act through related but distinct mechanisms—a lower substitution rate directly reduces genetic resolution by limiting the number of mutations that accumulate per transmission event, while a shorter incubation period compresses the time available for mutations to accrue before sampling, producing the same effect indirectly. Both push observed pairwise distances toward zero and concentrate them in a narrow range, eroding the signal on which any method must operate. This is consistent with the well-documented resolution ceiling in genomic epidemiology: when pathogen diversity is intrinsically low relative to the scale of transmission, no method can recover meaningful cluster structure from sequence data alone, regardless of how transmission uncertainty is modelled [5, 23, 24]. Losses compounded under parameter mismatch, but the degradation under matched conditions confirms that these are fundamental data limitations rather than inference artefacts.

Increased incubation CV revealed the clearest practical trade-off: under matched conditions most models improved, but under mismatched conditions deterministic-input models suffered large relative losses (F1 up to 25.4% for LD) while stochastic-input models were largely unaffected. This asymmetry reflects a structural difference in how each formulation handles variability—stochastic inputs absorb additional variance, whereas deterministic inputs encode it as systematic error [17, 34, 44, 45]. Combined with the baseline result, this defines a clear trade-off: deterministic inputs are stronger when assumptions hold, stochastic inputs when they do not. The robustness of LD and LS to substitution-rate perturbation suggests that supervised approaches may be preferable when clock rates are uncertain and labelled data are available. Removing clock relaxation consistently improved performance, suggesting that modelling rate variation adds noise when true evolutionary rate heterogeneity is minimal. Testing-delay misspecification had negligible effects throughout, reassuring given how poorly testing behaviour is often characterised in surveillance.

Several limitations deserve acknowledgement. EpiLink operates on consensus genomes and therefore ignores within-host diversity and minority variants that may carry additional transmission signal. The target scenario set S_*_ must be specified by the user; here, we limited it to direct transmission and co-primary infection, which is appropriate for recent linkage but would miss longer transmission chains. Compatibility scores are not calibrated posterior probabilities, and downstream clustering performance depends on the Leiden resolution parameter in ways that require careful sensitivity analysis, even though it remains generally robust to a broad range of permissible resolutions. Finally, the synthetic benchmark used a fixed transmission tree; performance on outbreak topologies with different heterogeneity profiles may differ.

Taken together, these results position EpiLink as most valuable when labelled training data are unavailable, when analysts need to explain why a pair or cluster has been flagged, or when the inferential target is recent linkage under superspreading conditions. In those settings, the explicit encoding of natural-history uncertainty into a graded compatibility score offers a principled and interpretable alternative to the ad hoc distance thresholds that remain dominant in routine surveillance.

## Supporting information

**S1 Text. Detailed derivation and modelling assumptions for the EpiLink compatibility model.** Provides the full derivation of the scenario-specific testing-time difference and transmission-related branch length used by EpiLink, together with the natural-history, observation, and molecular-clock assumptions.

**S1 Fig. Temporal stability of cluster partitions as the epidemic accumulates.** Forward overlap, backward overlap, and Jaccard index between consecutive weekly partitions, plotted against epidemic week for all six models. Stability is lowest in the first 1–2 epidemic weeks when cluster structure is sparse, rising rapidly, and remaining high thereafter. LS and ESD are the only models whose Jaccard index never falls below 0.7 across any transition.

**S2 Fig. Supplementary comparison with dated-tree TreeCluster on the synthetic baseline.** Best BCubed F1 for TreeCluster over method and threshold grid, compared with EpiLink-Leiden and logistic-regression-Leiden models. TreeCluster produced hard partitions only, so AP was not calculated.

**S3 Fig. Consistency of performance across perturbation scenarios.** Performance metrics under matched (inference parameters updated to reflect each perturbed data-generation scenario) and mismatched (inference parameters held fixed at baseline; data-generation parameters varied) conditions, evaluated across all 12 perturbation scenarios. The baseline performance ordering—LD strongest overall, ESD strongest EpiLink variant, and deterministic-input models outperforming stochastic-input counterparts—was broadly preserved across conditions. (A) Average precision (AP). (B) Best F1 score across Leiden resolution sweep. (C) Mean resolution stability (F1 between consecutive Leiden resolution steps; measures robustness to the choice of resolution parameter). (D) SD of resolution stability. Error bars show the 95% confidence interval.

**S4 Fig. Sensitivity of average precision (AP) to perturbations of data-generating parameters.**

Lollipop plot showing relative AP score loss compared with the unperturbed baseline for each model across twelve perturbation scenarios, under matched and mismatched inference conditions. Matched: inference parameters updated to reflect each perturbed scenario; mismatched: inference parameters held fixed at baseline. Each row corresponds to a perturbation scenario. Positive values indicate improved performance relative to baseline; negative values indicate degradation. Arrows indicate values exceeding the axis range.

**S5 Fig. Partition agreement between EpiLink-Leiden and dated-tree TreeCluster in the Boston dataset.** Adjusted Rand index and adjusted mutual information across TreeCluster avg clade thresholds and Leiden resolutions. The exported Boston TreeCluster partition used avg clade at 28 days.

## Data Availability

The EpiLink software is available under an MIT license at https://github.com/ydnkka/epilink and is archived on Zenodo at https://doi.org/10.5281/zenodo.20402076. All code, configuration files, and derived outputs required to reproduce the analyses presented in this manuscript are available in the evaluation branch at https://github.com/ydnkka/epilink/tree/evaluation and are archived on Zenodo at https://doi.org/10.5281/zenodo.20546213. The Boston empirical SARS-CoV-2 sequences and metadata were derived from the previously published Lemieux et al. dataset; assembled genomes and raw reads from that dataset are available through NCBI GenBank/SRA under BioProject PRJNA622837. The synthetic benchmark was derived from SCoVMod outputs described by Banks et al. No new individual-level clinical, genomic, or epidemiological data were collected for this study.

https://doi.org/10.5281/zenodo.20546213

https://github.com/ydnkka/epilink

https://doi.org/10.5281/zenodo.20402076

https://www.ncbi.nlm.nih.gov/bioproject/PRJNA622837

https://doi.org/10.1126/science.abe3261

https://github.com/Kao-Group/SCoVMod.git

https://doi.org/10.5281/zenodo.6420991

## Acknowledgments

The authors thank the researchers and public health teams who generated and shared the Boston SARS-CoV-2 genomic dataset reanalysed in this study.

