## Supplementary material for "EpiLink: a simulation-based compatibility model for genomic transmission clustering in infectious disease surveillance": S1 Text

### S1 Text. Detailed derivation and modelling assumptions for the EpiLink compatibility model.

This supplement expands the model summary in the main manuscript and makes explicit the structural assumptions underlying the default EpiLink score. For a sampled pair $(i, j)$, the observed data are a signed testing-time difference $t_{ij}$ (in days) and a consensus-level genetic distance $g_{ij}$ (in substitutions). EpiLink compares each observed pair against Monte Carlo draws generated under candidate latent transmission scenarios and reports a compatibility score, not a calibrated posterior probability.

#### Contents

**S1 Natural-history model** **1**

**S2 Transmission timing relative to infectiousness onset** **2**

**S3 Observation model for testing times** **2**

**S4 Scenario space** **3**

**S5 Temporal compatibility variable** **3**

**S6 Branch length and genetic model** **4**

**S7 Compatibility scoring** **5**

**S8 Modelling assumptions** **6**

**S9 Interpretation and scope** **7**

#### S1 Natural-history model

EpiLink uses an $E/P/I$ natural-history model in which the latent ($E$), presymptomatic infectious ($P$), and symptomatic infectious ($I$) stages have Gamma-distributed durations. The formulation follows the variable-infectiousness framework of Hart et al. [1] and is consistent with early SARS-CoV-2 evidence for substantial presymptomatic transmission and incubation periods centred near five days [3, 4, 5, 6].

Let

$$y_E \sim \Gamma(k_E, \theta_{\text{inc}}), \qquad y_P \sim \Gamma(k_P, \theta_{\text{inc}}), \qquad y_I \sim \Gamma(k_I, \theta_I),$$

where the presymptomatic shape is defined by

$$k_P = k_{\text{inc}} - k_E,$$

so that the incubation period is

$$\tau_{\text{inc}} = y_E + y_P \sim \Gamma(k_{\text{inc}}, \theta_{\text{inc}}).$$

In the current implementation, $k_I = 1$ and

$$\theta_I = \frac{1}{k_I \mu} = \frac{1}{\mu},$$

where $\mu$ is the symptomatic-stage removal rate. The constraint $0 < k_E < k_{\text{inc}}$ ensures a positive presymptomatic stage duration.

#### S2 Transmission timing relative to infectiousness onset

Let $y_{\text{toit}}$ denote the time from onset of infectiousness to transmission. In the implementation, $y_{\text{toit}}$ is drawn from the `InfectiousnessToTransmission` profile, whose density is [1]

$$f(y_{\text{toit}}) = \begin{cases} 0, & y_{\text{toit}} < 0, \\ C\left[\alpha(1 - F_P(y_{\text{toit}})) + \int_0^{y_{\text{toit}}} (1 - F_I(y_{\text{toit}} - y_P)) f_P(y_P)\, dy_P\right], & y_{\text{toit}} \geq 0, \end{cases}$$

where $f_P$ and $F_P$ are the presymptomatic probability density and cumulative distribution functions, $F_I$ is the symptomatic cumulative distribution function, and $\alpha$ is the presymptomatic-to-symptomatic transmission-rate ratio.

Writing $\lambda_{\text{inc}} = 1/(k_{\text{inc}}\theta_{\text{inc}})$, the Hart et al. normalisation constant is

$$C = \frac{k_{\text{inc}}\lambda_{\text{inc}}\mu}{\alpha k_P \mu + k_{\text{inc}}\lambda_{\text{inc}}}.$$

The implied presymptomatic transmission fraction is

$$q_P = \frac{\alpha k_P \mu}{\alpha k_P \mu + k_{\text{inc}}\lambda_{\text{inc}}}.$$

Because $\alpha > 1$ under the default parameterisation, transmission intensity is elevated before symptom onset and reduced after, although symptomatic transmission remains possible. This is consistent with empirical evidence that infectiousness peaks around symptom onset and that a substantial fraction of SARS-CoV-2 transmission occurs before symptoms are recognised [3, 4].

The generation interval for a single infection event is then

$$\tau_{\text{gen}} = y_E + y_{\text{toit}},$$

broadly consistent with empirical estimates from transmission-pair data [7, 2].

For the default Hart et al. 2021 parameterisation used in EpiLink, the natural-history parameters are

$$k_{\text{inc}} = 5.807, \qquad \theta_{\text{inc}} = 0.948 \text{ days}, \qquad k_E = 3.38, \qquad k_I = 1,$$

$$\mu = 0.37 \text{ day}^{-1}, \qquad \alpha = 2.29.$$

These imply $k_P = k_{\text{inc}} - k_E = 2.427$, a mean incubation period of approximately 5.51 days, and a mean symptomatic-stage duration of approximately 2.70 days.

#### S3 Observation model for testing times

Observed case times are sampling or testing times, not infection times. EpiLink assumes a Gamma-distributed delay from symptom onset to testing,

$$x_{\text{test}} \sim \Gamma(k_{\text{test}}, \theta_{\text{test}}),$$

drawn independently for each sampled case. The total delay from infection to testing is therefore

$$\tau_{\text{obs}} = \tau_{\text{inc}} + x_{\text{test}}.$$

For a latent reference infection time $t_x$ and a scenario-specific elapsed time $A_k(s)$ from that reference point to the infection of sampled case $k \in \{i, j\}$,

$$t_{\text{test},k} = t_x + A_k(s) + \tau_{\text{obs},k}.$$

The baseline testing-delay parameters used in this study are mean = 1.0 day and coefficient of variation = 1.0 (i.e. exponential), giving $k_{\text{test}} = 1$ and $\theta_{\text{test}} = 1.0$ day.

#### S4 Scenario space

Let $M$ denote the maximum hidden depth considered between the two sampled cases. The full candidate scenario set is

$$\mathcal{S}_M = \{H_{\text{AD}}(m) : m = 0, \ldots, M\} \cup \{H_{\text{CA}}(m_i, m_j) : m_i, m_j \geq 0,\ m_i + m_j \leq M\}.$$

Here:

- $H_{\text{AD}}(m)$ is an ancestor-descendant history in which case $j$ descends from case $i$ through $m$ unsampled intermediates;
- $H_{\text{CA}}(m_i, m_j)$ is a common-ancestor history in which both sampled cases descend from a shared unsampled source through branch depths $m_i$ and $m_j$.

The labels $H_{\text{CA}}(m_i, m_j)$ and $H_{\text{CA}}(m_j, m_i)$ are generally distinct because EpiLink uses the signed time difference $t_{ij}$. The default target used throughout this manuscript is intentionally narrow:

$$\mathcal{S}_\star = \{H_{\text{AD}}(0),\ H_{\text{CA}}(0, 0)\}, \qquad M = 0.$$

The reported target score is therefore the sum of compatibility with direct transmission ($H_{\text{AD}}(0)$, $i$ directly infects $j$) and co-primary infection ($H_{\text{CA}}(0, 0)$, both sampled cases descend from the same unsampled source in a single generation step). Note that $H_{\text{AD}}(0)$ and $H_{\text{CA}}(0, 0)$ target different directions of time: the ancestor-descendant scenario implies $j$ is later than $i$ (positive $t_{ij}$), while the common-ancestor scenario is symmetric in timing.

#### S5 Temporal compatibility variable

Subtracting the testing times for cases $i$ and $j$ gives the scenario-implied testing-time difference

$$T_s = A_j(s) - A_i(s) + \tau_{\text{obs},j} - \tau_{\text{obs},i}.$$

For ancestor-descendant scenarios, the index case $i$ is the reference point ($A_i = 0$):

$$A_i(H_{\text{AD}}(m)) = 0, \qquad A_j(H_{\text{AD}}(m)) = \sum_{r=0}^{m} \tau_{\text{gen},r},$$

so

$$T_{ij}(H_{\text{AD}}(m)) = \sum_{r=0}^{m} \tau_{\text{gen},r} + \tau_{\text{obs},j} - \tau_{\text{obs},i}.$$

For common-ancestor scenarios, both sampled cases descend from an unsampled source (the reference point at elapsed time 0):

$$A_i(H_{\text{CA}}(m_i, m_j)) = \sum_{r=1}^{m_i+1} \tau^{(i)}_{\text{gen},r}, \qquad A_j(H_{\text{CA}}(m_i, m_j)) = \sum_{s=1}^{m_j+1} \tau^{(j)}_{\text{gen},s},$$

giving

$$T_{ij}(H_{\mathrm{CA}}(m_i,m_j)) = \sum_{s=1}^{m_j+1} \tau_{\mathrm{gen},s}^{(j)} - \sum_{r=1}^{m_i+1} \tau_{\mathrm{gen},r}^{(i)} + \tau_{\mathrm{obs},j} - \tau_{\mathrm{obs},i}.$$

Monte Carlo simulation under scenario $s$ with $N = 10{,}000$ draws yields the empirical temporal distribution $\{T_s^{(1)}, \ldots, T_s^{(N)}\}$.

### S6 Branch length and genetic model

EpiLink converts latent transmission histories into an effective transmission-related branch length $B_s$ (measured in days), and then into a distribution of expected consensus genetic distances.

#### S6.1 Ancestor-descendant branch length

For ancestor-descendant scenarios:

$$\widetilde{B}_{ij}(H_{\mathrm{AD}}(m)) = \sum_{r=0}^{m} \tau_{\mathrm{gen},r} + \tau_{\mathrm{obs},j} - \tau_{\mathrm{obs},i}.$$

This equals the temporal contrast $T_{ij}(H_{\mathrm{AD}}(m))$: ancestor-descendant genetic separation is driven by the same elapsed time that determines the expected testing gap. Because lineage lengths cannot be negative, the implementation uses

$$B_{ij}(H_{\mathrm{AD}}(m)) = \max\{0,\ \widetilde{B}_{ij}(H_{\mathrm{AD}}(m))\}.$$

#### S6.2 Common-ancestor branch length

For common-ancestor scenarios, branch length accumulates along both descendant lineages from the shared unsampled source:

$$B_{ij}(H_{\mathrm{CA}}(m_i,m_j)) = \sum_{r=1}^{m_i+1} \tau_{\mathrm{gen},r}^{(i)} + \sum_{s=1}^{m_j+1} \tau_{\mathrm{gen},s}^{(j)} + \tau_{\mathrm{obs},i} + \tau_{\mathrm{obs},j}.$$

Unlike the ancestor-descendant case, this sum is always non-negative and grows with each additional unsampled intermediate.

#### S6.3 Molecular clock

Let $\nu$ denote the median substitution rate in substitutions per site per year and $L$ the genome length in sites. The per-genome daily rate is

$$\lambda = \frac{\nu L}{365}.$$

Under a strict molecular clock, $\lambda$ is fixed across draws. Under the relaxed-clock option, EpiLink samples branch-specific rates from an uncorrelated log-normal model [8],

$$\nu^{(n)} \sim \mathrm{LogNormal}\big(\log \nu - \tfrac{1}{2}\sigma_\nu^2,\ \sigma_\nu^2\big), \qquad \lambda^{(n)} = \frac{\nu^{(n)} L}{365},$$

where $\sigma_\nu$ is the clock-relaxation parameter and the log-normal is parameterised so that the *median* equals $\nu$. The baseline evaluation uses $\nu = 10^{-3}$ substitutions per site per year and $\sigma_\nu = 0.33$ for the relaxed-clock variant.

##### S6.4 Mutation model

Given a branch-length draw $B_s^{(n)}$, the expected genetic distance is

$$G_s^{(n)} = \lambda^{(n)} B_s^{(n)}$$

under the *deterministic* mutation model, or

$$G_s^{(n)} \mid B_s^{(n)}, \lambda^{(n)} \sim \text{Poisson}(\lambda^{(n)} B_s^{(n)})$$

under the *stochastic* mutation model. The deterministic model gives a single expected substitution count given branch length and clock rate; the stochastic model adds Poisson noise around that expectation, substantially widening the compatibility region near the origin of the temporal-genetic score surface (see main text, Results).

#### S7 Compatibility scoring

For each scenario $s$, EpiLink caches Monte Carlo draws

$$\{T_s^{(n)},\ B_s^{(n)},\ G_s^{(n)}\}_{n=1}^{N}.$$

Observed values are scored by their percentile within the simulated distribution. For an observed value $x_{\text{obs}}$ and $N$ scenario-specific draws $x_1^{(s)}, \ldots, x_N^{(s)}$,

$$p_s(x_{\text{obs}}) = \frac{\#\{n : x_n^{(s)} \le x_{\text{obs}}\}}{N},$$

and the percentile is converted to a centrality-based compatibility score:

$$C_s(x_{\text{obs}}) = 1 - 2|p_s(x_{\text{obs}}) - 0.5|.$$

This transformation maps percentile $p_s$ to a score that is 1 at the median of the scenario distribution and declines symmetrically towards 0 at the extremes. Applying this to time and genetics gives temporal and genetic compatibilities,

$$C_{T,s}(i,j) = C_s(t_{ij}), \qquad C_{G,s}(i,j) = C_s(g_{ij}),$$

and the scenario-level compatibility is their product:

$$C_s(i,j) = C_{T,s}(i,j)\, C_{G,s}(i,j).$$

The multiplicative combination rewards pairs that are simultaneously central in both the temporal and genetic distributions implied by the same latent scenario. For a user-defined target subset $\mathcal{S}_\star \subseteq \mathcal{S}_M$, EpiLink reports the summed target score:

$$C_{\mathcal{S}_\star}(i,j) = \sum_{s \in \mathcal{S}_\star} C_s(i,j).$$

Because these are summed compatibilities, not probabilities, target scores can exceed 1 when both scenarios simultaneously align well with the observation (as occurs near the common-ancestor / direct-transmission overlap region at small $t_{ij}$ and $g_{ij}$).

#### S8 Modelling assumptions

The derivation above makes the following assumptions, which are useful to state explicitly.

1. **Recent linkage is represented by a restricted latent-history set.** In this manuscript, "recent transmission linkage" is defined as either direct transmission or co-primary infection from a shared recent source. Unsampled intermediates are not part of the default target score because $M = 0$.

2. **Individuals share a common natural-history model.** Stage-duration distributions, testing-delay distributions, and infectiousness parameters are assumed identical across all cases within a run of the model. Individual-level heterogeneity is not accounted for.

3. **Transmission timing is anchored to symptom onset.** The $E/P/I$ profile assumes higher transmission intensity in the presymptomatic phase and a reduced propensity after symptom onset, rather than a flat infectiousness profile over time [1, 3, 4].

4. **Testing time is a noisy proxy for infection time.** The only observed time used by EpiLink is the sample collection or testing date. Delays from symptom onset to testing are treated as independent Gamma-distributed random variables and are not directly observed.

5. **Generation intervals are independent and identically distributed across transmission events.** Once the natural-history parameters are fixed, each transmission event contributes an independent draw from the same generation-interval distribution. Correlations arising from, for example, household or healthcare transmission contexts are not modelled.

6. **Consensus divergence is driven primarily by transmission-linked lineage time.** EpiLink assumes limited within-host diversity over the timescale of interest, so that consensus differences mostly accumulate along the between-host lineages connecting the sampled genomes. This is broadly consistent with evidence for low within-host diversity at high viral load and a narrow SARS-CoV-2 transmission bottleneck [9].

7. **Mutation counts are conditionally independent given branch length and clock rate.** Under the stochastic mutation model, the observed consensus distance is generated as a Poisson count conditional on the effective branch length and the (possibly relaxed-clock-drawn) substitution rate. Spatial or phylogenetic correlations in substitution rates are not modelled.

8. **Temporal and genetic compatibility are combined multiplicatively.** The product $C_s(i,j) = C_{T,s}(i,j) \cdot C_{G,s}(i,j)$ rewards pairs that are simultaneously central in both the temporal and genetic distributions implied by the same latent scenario, and penalises pairs that are extreme in either dimension. An alternative additive or max-based aggregation would assign different weights to these two evidence streams.

9. **Compatibility is not a posterior probability.** The percentile-centrality score is intentionally descriptive and symmetric about the scenario median. A score of 0 indicates that the observed value falls at the extreme of the Monte Carlo distribution under that scenario; a score of 1 indicates it falls exactly at the median. Low compatibility indicates poor alignment with the chosen assumptions; it does not by itself prove the absence of epidemiological linkage or provide a calibrated false-positive rate.

#### S9 Interpretation and scope

Under this formulation, EpiLink should be read as an interpretable compatibility filter for recent-linkage hypotheses. Its outputs depend directly on the chosen latent-history target $\mathcal{S}_\star$, the assumed natural-history profile, the testing-delay model, and the molecular clock model. In the present study the target is restricted to $\{H_{\mathrm{AD}}(0), H_{\mathrm{CA}}(0,0)\}$; expanding the target to include hidden intermediates (larger $M$) would assign positive compatibility to pairs connected through unsampled chains, broadening sensitivity at the cost of reduced specificity for recent direct linkage.

The method is most useful when the practical goal is to rank or cluster plausible recent-transmission neighbourhoods rather than estimate a fully calibrated transmission tree, consensus genomes and sampling dates are available, labelled training pairs are absent or cannot be assumed to transfer across settings, and the analyst can inspect the natural-history and molecular-clock assumptions encoded in the model parameters. Because each score decomposes into the temporal and genetic compatibility contributions from each target scenario, it is possible to diagnose directly why a particular pair was or was not flagged as a recent-linkage candidate.
