## Supplementary figures and images for "EpiLink: a simulation-based compatibility model for genomic transmission clustering in infectious disease surveillance"

### S1 Fig

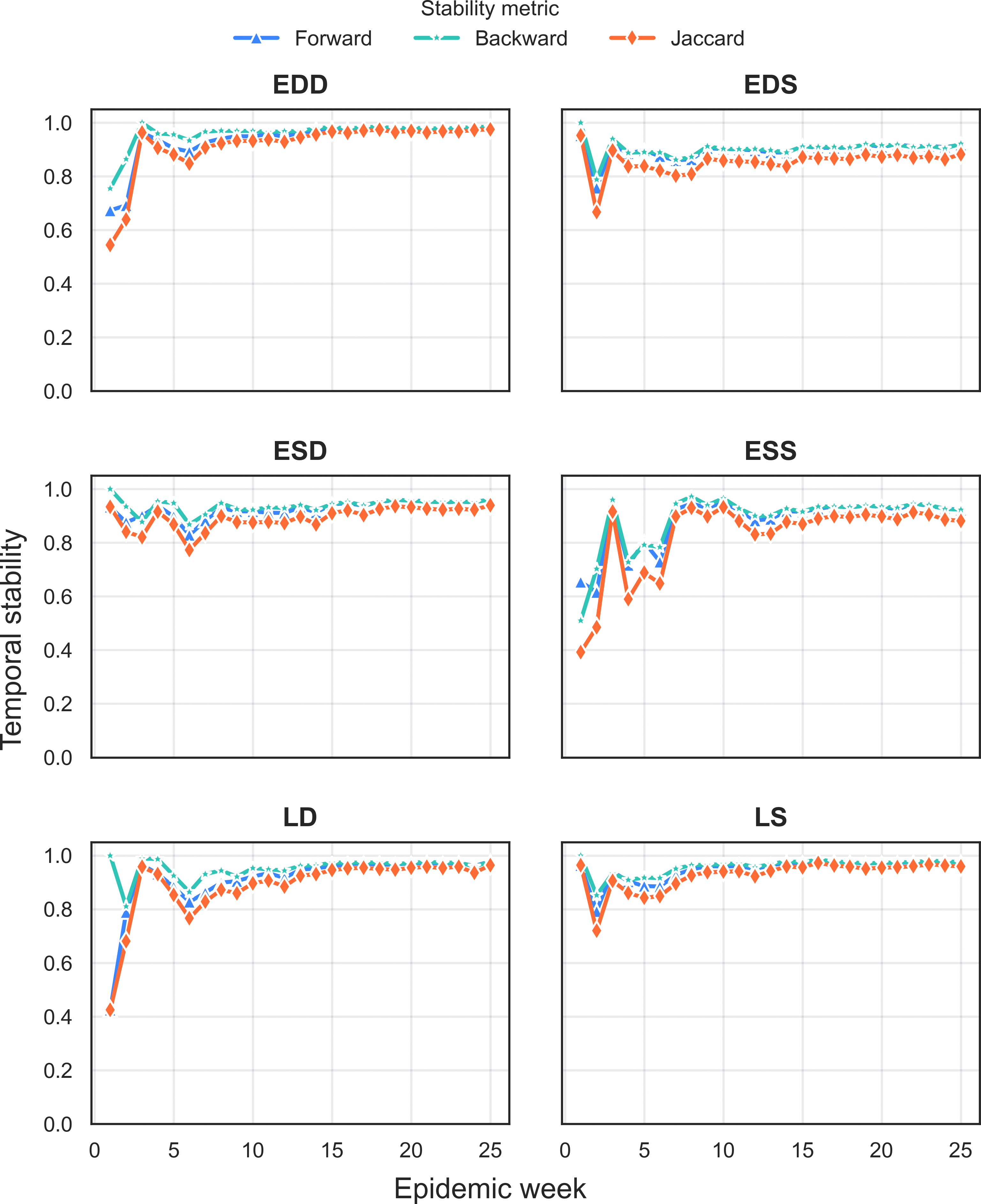

### S2 Fig

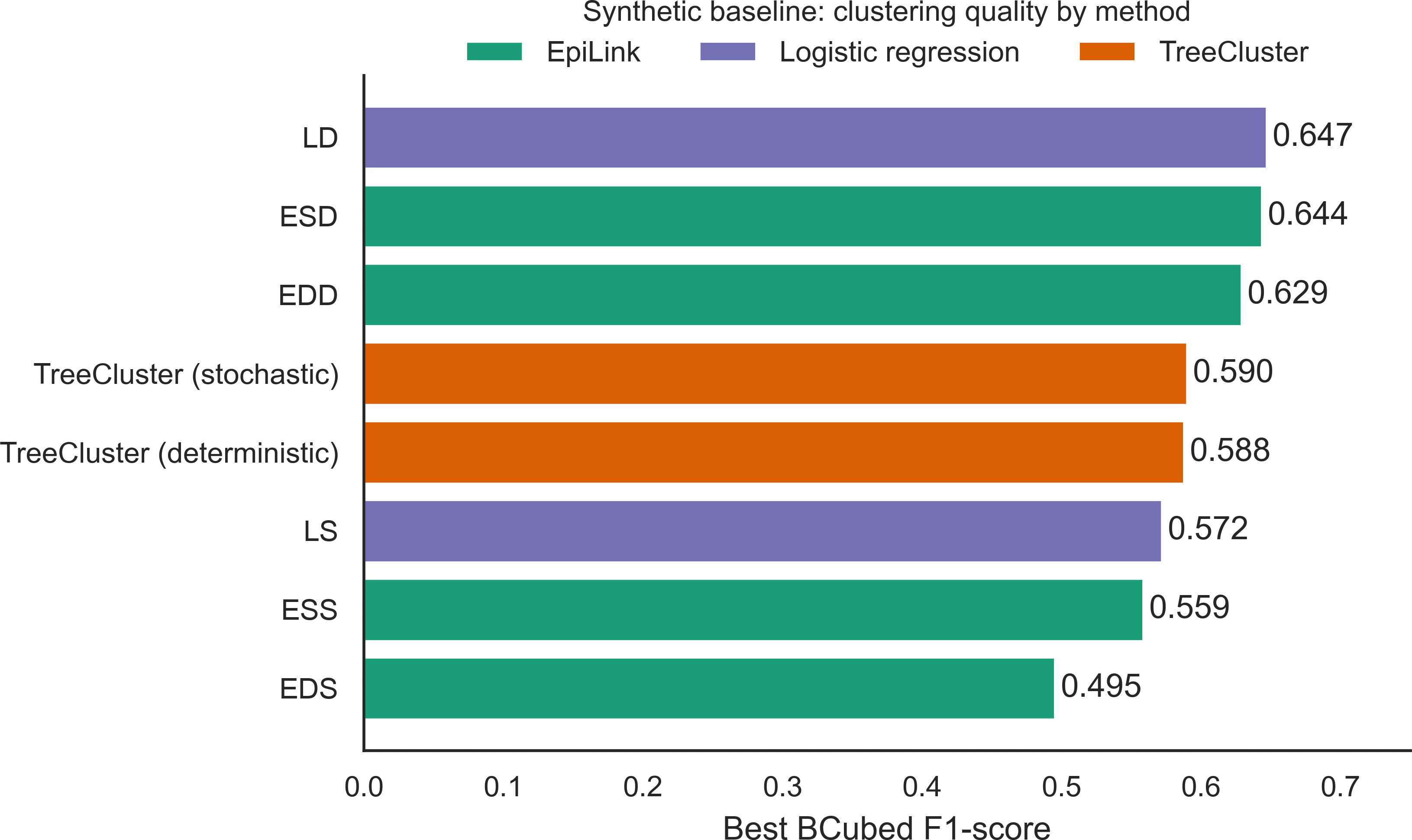

### S3 Fig

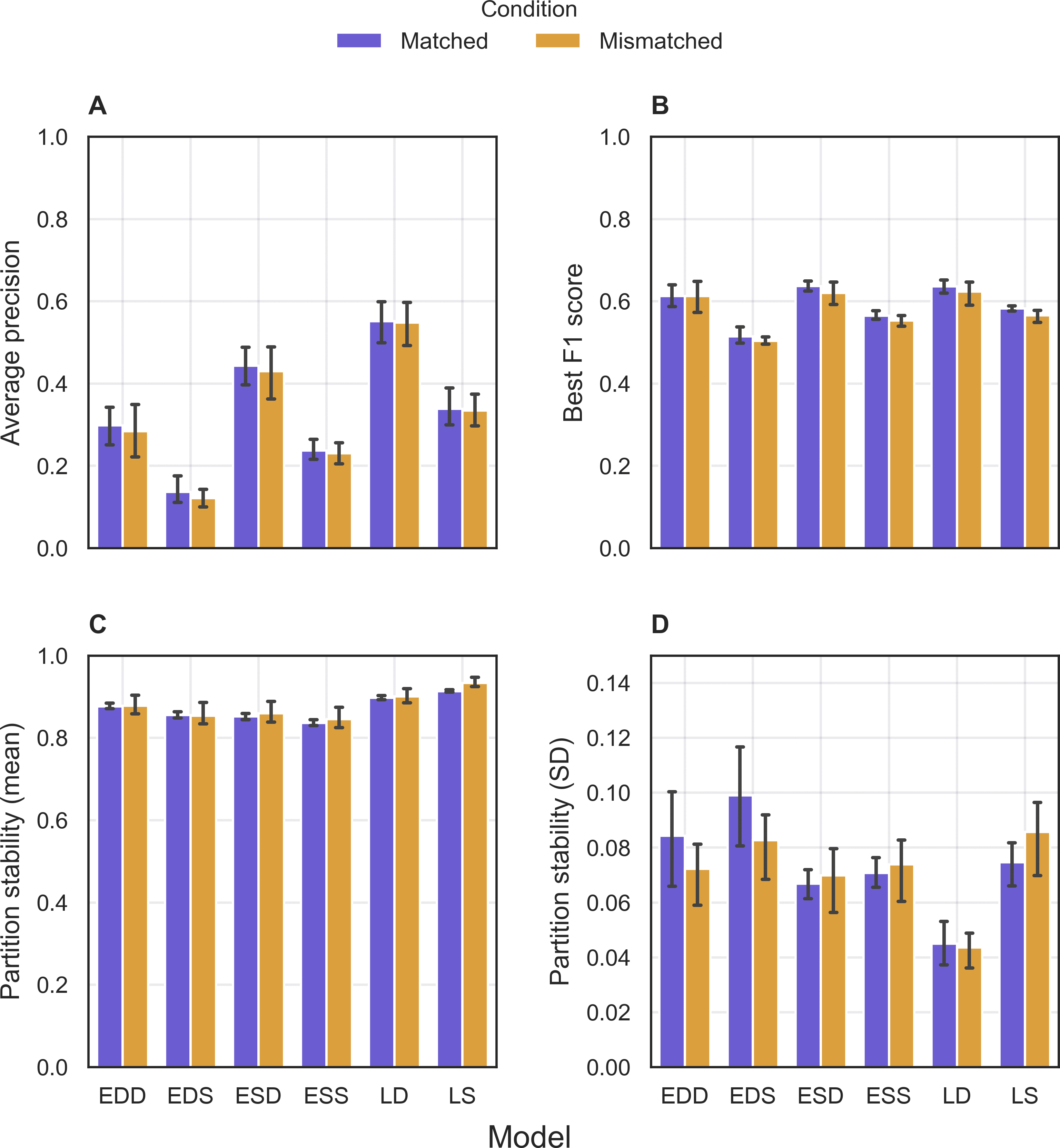

### S4 Fig

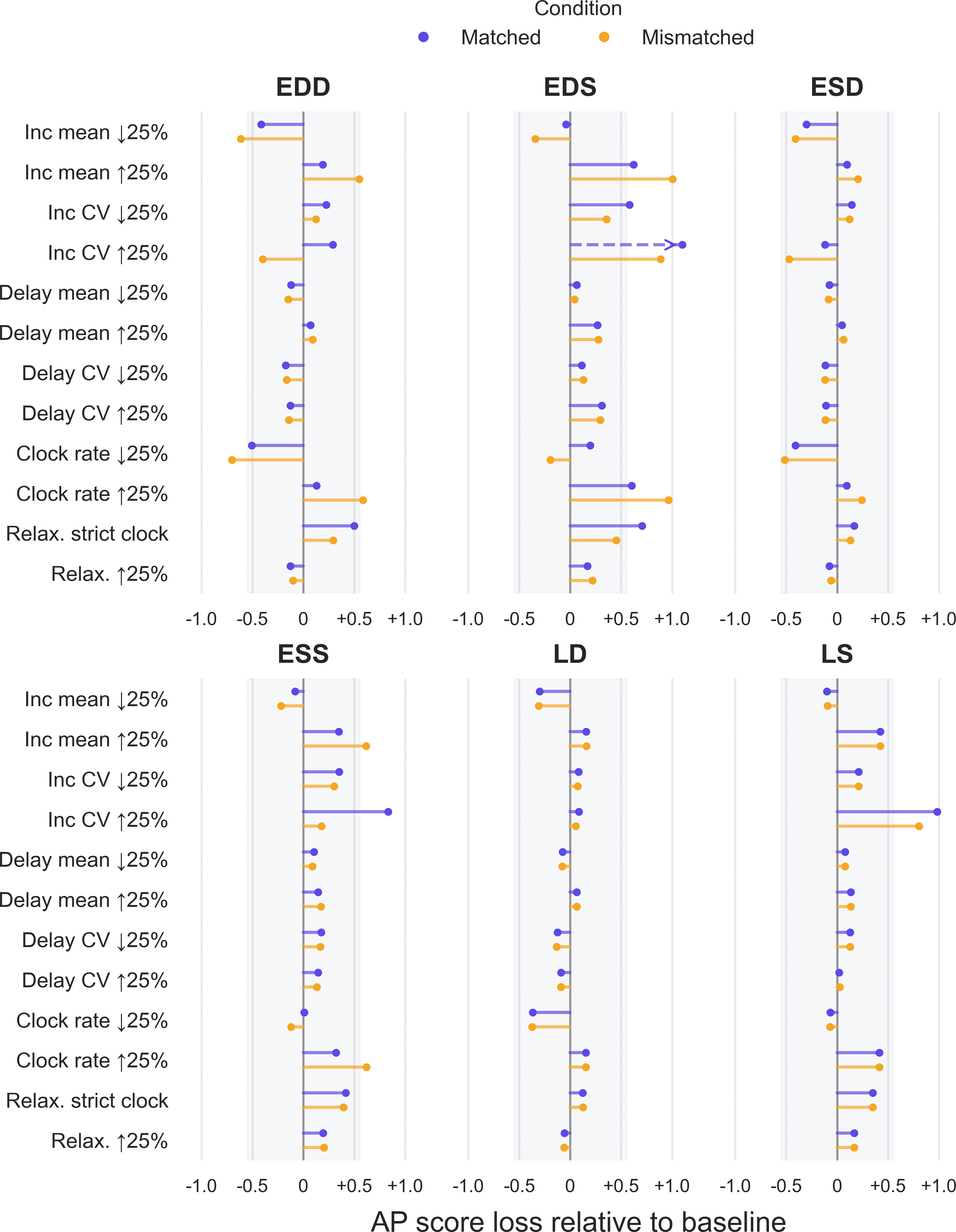

### S5 Fig

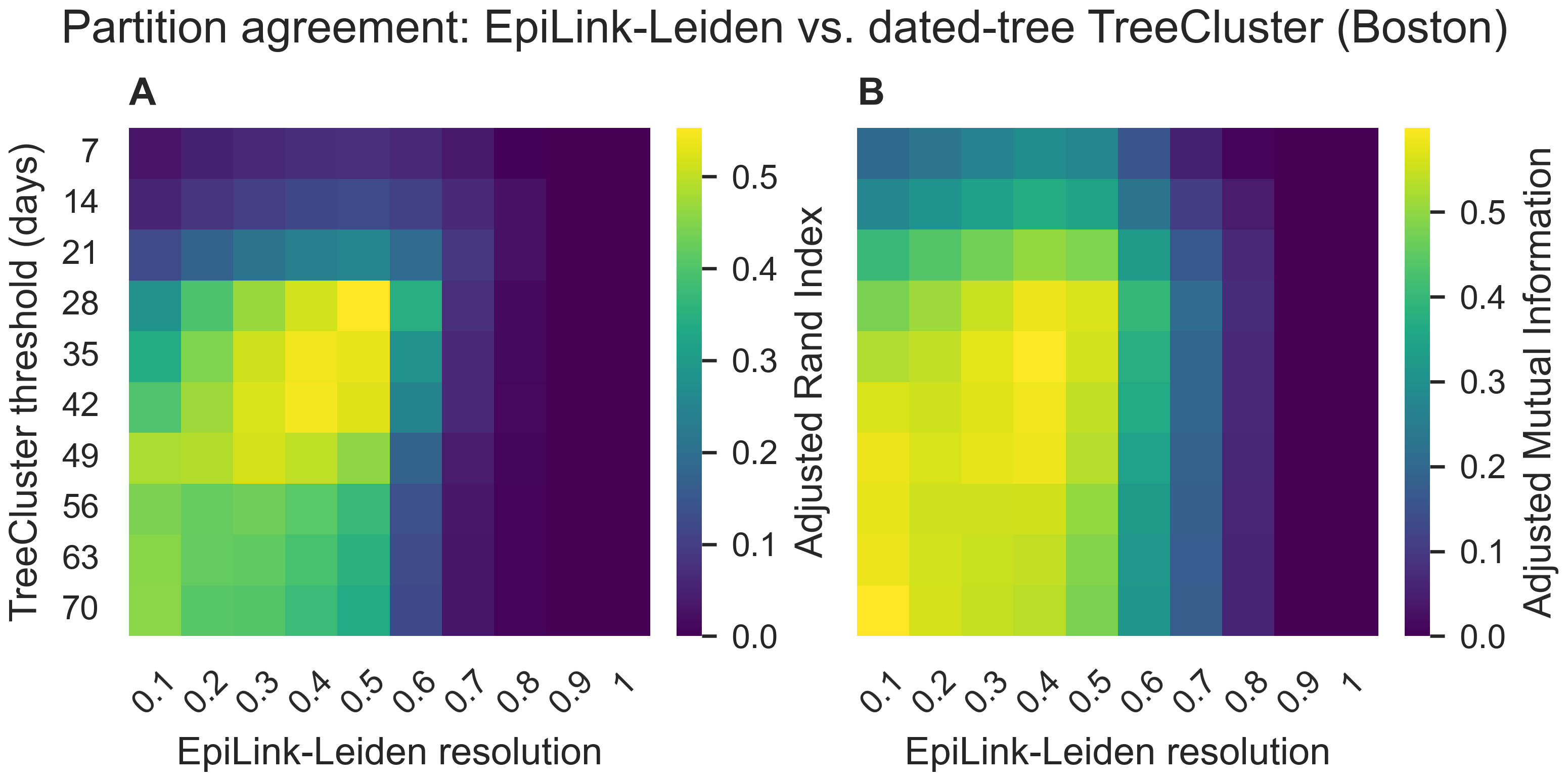
